# Health impacts of takeaway management zones around schools in six different local authorities across England: a public health modelling study using PRIMEtime

**DOI:** 10.1101/2024.06.11.24308755

**Authors:** Nina Trivedy Rogers, Ben Amies-Cull, Jean Adams, Michael Chang, Steven Cummins, Daniel Derbyshire, Suzan Hassan, Matthew Keeble, Bochu Liu, Antonieta Medina-Lara, Bea Savory, John Rahilly, Richard Smith, Clare Thompson, Martin White, Oliver Mytton, Thomas Burgoine

**Author notes:** Corresponding author: Nina T Rogers, MRC Epidemiology Unit, University of Cambridge School of Clinical Medicine, Box 285 Institute of Metabolic Science, Cambridge Biomedical Campus, Cambridge, CB2 0QQ, UK.

## Abstract

**Background:** In England, the number of takeaway food outlets (‘takeaways’) has been increasing for over two decades. Takeaway management zones around schools are an effective way to restrict the growth of new takeaways but their impacts on population health have not been estimated.

**Methods:** To model the impact of takeaway management zones on health, we used estimates of change in and exposure to takeaway outlets (across home, work, and commuting buffers) based on a previous evaluation suggesting that 50% of new outlets were prevented from opening because of management zones. Based on previous cross-sectional findings, we used changes in takeaway exposure to estimate changes in BMI, from 2018 to 2040. Finally, we used PRIMEtime, a proportional multistate lifetable model, and BMI change to estimate the impact of the intervention, in a closed-cohort of adults (25-64 years), in terms of incidence of 12 non-communicable diseases, obesity prevalence, quality-adjusted life years (QALYs) and healthcare costs saved by 2040 in six selected local authorities across the rural-urban spectrum in England (Wandsworth, Manchester, Blackburn with Darwen, Sheffield, North Somerset, and Fenland).

**Results:** By 2031, compared to no intervention, reductions in outlet exposure ranged from 3 outlets/person in Fenland to 28 outlets/person in Manchester. This corresponded to per person reductions in BMI of 0.68 and 0.08 kg/m^2^, respectively. Relative to no intervention, obesity prevalence was estimated to be reduced in both sexes in all LAs, including by 2.3 percentage points (PP) (95% uncertainty interval:2.9PP, 1.7PP) to 1.5PP (95%UI:1.9PP, 1.1PP) in males living in Manchester and Wandsworth by 2040, respectively. Model estimates showed reductions in incidence of disease, including type II diabetes (eg: 964 (95%UI:1565, 870) fewer cases /100,000 population for males in Manchester)), cardiovascular diseases, asthma, certain cancers and low back pain. Savings in healthcare costs (millions(£)) ranged from £0.90 (95%UI: £1,23, £0.54) in Fenland to £5.44 (95%UI:£3.87, £7.45) in Manchester. Gains in QALYs/100,000 person were broadly similar across local authorities.

**Conclusions:** Takeaway management zones in England have the potential to meaningfully contribute towards reducing obesity prevalence and associated healthcare burden in the adult population, both at the local level and across the rural-urban spectrum.

## Introduction

Meals purchased out-of-home, including foods from takeaway food outlets (“takeaways”), are typically energy dense and high in sugar and salt, but low in micronutrients, and tend to be served in large portions^1–3^. Consumption of takeaway food is associated with lower diet quality, higher energy intake and body mass index (BMI), weight gain and greater risk of obesity^4,5^. This may be a result of passive over-consumption of takeaway foods, which bypass regular human satiety mechanisms^6^. In turn, poor diet and excess weight are risk factors for diseases including type II diabetes and cardiovascular disease^7–9^.

Neighbourhood food environments have become a focus for public health action as they may encourage unhealthy dietary behaviours ^10^. Local residential exposure to takeaways has been associated with higher levels of takeaway food consumption, BMI and risk of obesity in adults^11–14^ and children^15^ although this relationship has not been observed in all studies^16^. Differences between takeaway exposure and change in BMI have been hypothesised to differ between neighbourhoods in urban and rural areas due to differences in the structure of the built environment, and in the dietary patterns and levels of overweight of residents^17,18^. However, one study in the Netherlands found that takeaway exposure within 1km of the home was associated with higher BMI in both rural and urban populations^19^.

In the UK, takeaways continue to increase in number, with 47,961 registered in 2023, equating to a 2.8% increase per year from 2018 onwards^20^. In one study in Norfolk, England, longer term data suggests that the density of takeaways increased by approximately 44% over an 18 year period from 1990 to 2008, with higher densities and stronger growth in more deprived areas^21^. A 34% increase in expenditure on takeaway food from £7.9 billion in 2009 to £9.9 billion in 2016 has previously been reported by the takeaway food industry^22^.

Takeaways have been shown to cluster within walking distance of schools in England and other countries^23,24^. In England, and often with the stated intention of improving health, urban planners can use existing powers to prevent new takeaways opening, thereby limiting growth in people’s future exposure to takeaways. These “takeaway management zones” are commonly centred on schools, for example where no new takeaways are permitted within 400m radius of a school site. These are also sometimes referred to by local authorities (LAs) as takeaway “exclusion zones”. It has previously been estimated that management zones in England covered an average of 17% of land area in the LAs in which they have been adopted; a significant spatial footprint with capacity therefore to affect whole populations, in addition to children^25^. A recent study showed that implementation of management zones was associated with a 54% reduction in the number of new takeaways at up to six years post-intervention^26^. This is likely due to a combination of a decrease in the number of planning applications submitted for new takeaways, and an increase in the percentage of these applications being rejected, which was also observed in these areas^25^. However, the extent to which takeaway management zones around schools may benefit population health due to this retail change has not been explored.

Evaluating the future health impacts of takeaway management zones around schools is important to inform uptake and implementation. A lack of evidence in this regard has been cited as a barrier to adoption ^27,28^. Such evidence is also important in the defence of takeaway management zones against legal challenges as the proportionality principle requires potential harms to private interests be offset by the likelihood of benefits to the public^29^. However, as the future is uncertain this is inherently difficult. The potential positive health impacts of takeaway management zones may also accrue over a long time-period, making it challenging and untimely to *observe* the effects of policy adoption. However, mathematical modelling can be used to predict future impacts and help inform decision making^30^. The PRIMEtime model is a multistate lifetable that has been used to estimate the health impacts of other interventions such as the UK soft drink industry levy and restrictions of television advertising unhealthy foods to children^31^. In this study, we aimed to estimate the future health impacts, to 2040, of the adoption of takeaway management zones around schools in six different LAs across England.

## Methods

### Scenarios of restricted future takeaway growth

We used data from a previously published forecast model to the year 2031 of mean changes in population exposure to takeaways in absence of the intervention i.e. under business-as-usual conditions. Briefly, the model used historical observed rates of growth in takeaways in non-adopter LAs that were similar in terms of urban-rural class to six purposively selected LAs: Wandsworth, Manchester, Sheffield, Blackburn with Darwen, North Somerset and Fenland (Table 1)^32^. These LAs were selected to represent classes across the rural-urban spectrum and to ensure geographical breadth across England. The selection also represented LAs that were either adopters of management zones around a similar year (Wandsworth, Manchester and Blackburn with Darwen) or hypothetical adopters (Sheffield, North Somerset and Fenland). Consequently, we also focus on these six LAs in our analysis. Population exposure to takeaways within LAs was measured across home, work and commuting domains, using census travel to work data^32^. Exposure to takeaways has previously been measured across these same three domains^33^. Full details of this forecast and population exposure model have been published elsewhere^31^.

**Table 1:** Demographic and urban-rural characteristics of six specified local authorities.

| Local Authority | Rural Urban Classification | Income Deprivation Quintile<sup>1</sup> | Population density<sup>2</sup> | Gender of adult population aged 25-64 years (N)<sup>3</sup> |  |  | Age group, N (%) |  |
| --- | --- | --- | --- | --- | --- | --- | --- | --- |
|  |  |  |  | Males | Female | All | 25-44 years | 45-64 years |
| Wandsworth | London urban with major conurbation | 3 | 9560 | 99,161 | 106,709 | 205,870 | 142,344 (69.1) | 63,526 (30.9) |
| Manchester | Urban with major conurbation (non-London) | 1 | 4773 | 149,682 | 139,056 | 288,738 | 187,934 (65.1) | 100,804 (34.9) |
| Sheffield | Urban with minor conurbation | 2 | 1513 | 145,487 | 145,249 | 290,736 | 157,524 (54.2) | 133,212 (45.8) |
| Blackburn with Darwen | Urban with city and town | 1 | 1129 | 38,110 | 37,792 | 75,902 | 38,110 (50.2) | 37,792 (49.8) |
| North Somerset | Urban with significant rural | 3 | 580 | 133,990 | 140,745 | 274,735 | 117,521 (42.8) | 157,214 (57.2) |
| Fenland | Largely or mainly rural | 2 | 188 | 25,282 | 25,583 | 50,865 | 23,405 (46.0) | 27,460 (54.0) |

<sup>1</sup> Income Deprivation quintile is for the whole population. Most deprived quintile = 1
(see: https://www.ons.gov.uk/peoplepopulationandcommunity/personalandhouseholdfinances/incomeandwealth/datasets/mappingincomedeprivationatlocalauthoritylevel)
<sup>2</sup> Population/square km (in relation to the whole population) https://www.ons.gov.uk/datasets/TS006/editions/2021/versions/4?f=get-data
<sup>3</sup> Numbers are based on PRIMEtime data in 2015

In this study, relative to business-as-usual growth, we modelled impacts of policy adoption under a realistic scenario where there was a 50% reduction in new takeaways, informed by previous research^26^. While takeaway management zones were adopted between 2015 and 2017, we aligned implementation dates to 2018 to allow for comparison between LAs. We also carried out sensitivity analysis under perfect implementation scenarios, whereby there was a100% reduction in new takeaways following the intervention. We assumed the policy was in place between 2018 and 2031 but given that forecasting in the longer term may lead to less precise estimates we assumed that any differences between business as usual and the intervention remained constant thereafter to 2040. Estimation of lower and upper confidence intervals for the three interventions were performed in R version 4.1.0.

### Relationship between change in takeaway exposure and BMI

In a previous study of UK adults aged 29-62 years, those most exposed (quartile 4) to takeaways across home, work and commuting domains had on average 1.21 kg/m^2^ (95% CI 0.68, 1.74) greater BMI than those least exposed (quartile 1)^33^. This is equivalent to an increase in BMI of 0.0241 kg/m^2^ for each additional takeaway a person is exposed to on a regular basis (unpublished results). Similar results from other analyses of these same data, but accounting only for home and work takeaway exposure, have also been reported^11^. This magnitude of association was similar to findings from a separate study using data from the Fenland Study, which showed 0.14 kg/m^2^ higher BMI per five additional takeaways exposed to^34^. We used this figure to estimate mean change in BMI attributable to change in per person exposure to takeaways within each LA for adults aged 25-64 years. In supplementary analysis we estimated mean population BMI change across quintiles of deprivation within LAs (see Table S3) using estimates of takeaway exposure change and the same value for the relationship between takeaway exposure change and BMI.

### Health impact modelling using PRIMEtime

We used PRIMEtime, a proportional multistate lifetable model, to simulate the impact of observed changes in BMI on a range of diet-related chronic diseases and other health outcomes. The PRIMEtime model works by simulating a change in obesity prevalence attributable to the intervention. It then estimates changes in incidence of specified BMI-related diseases and in disease-specific death rates while keeping deaths unrelated to obesity stable. In our main analysis we estimated the health impacts for a closed cohort of adults aged 25-64 years across 22 years (2018-2040) for each of the six LAs, assuming realistic implementation. We used Microsoft Excel to conduct 1000 runs of a Monte Carlo analysis in PRIMEtime, to estimate lower and upper uncertainty intervals (UI) of cases for 12 BMI-related non-communicable diseases and their associated quality adjusted life years (QALYs) benefits and healthcare cost saving outcomes.

Diseases related to BMI that were modelled in PRIMEtime were type II diabetes, ischaemic heart disease (IHD), atrial fibrillation/flutter, stroke, hypertensive heart disease, asthma, colon and rectum cancer, oesophageal cancer, breast cancer (females only), osteoarthritis (hip and knee) and low back pain. For estimating healthcare costs in PRIMEtime, disease-specific costs for each modelled disease are based on a range of routine national datasets including hospital episodes statistics admissions data, furthermore a detailed description of the model, including how healthcare costs are attributed to disease burden has been published previously^35^. QALYs were also estimated using utility weights and discounted using published National Institute for Health and Care Excellence (NICE) rates at a flat 3.5% for all health and costs^36^. In our results section we show total healthcare cost savings and QALYs gained in specific LAs and we also adjust the values by dividing them by the number of adults aged 25-64 living in a specific LA in 2018 and then multiplying by 100,000 to show values per 100,000 population. To ensure relevance of our estimates (i.e. because the associations between takeaway exposure and BMI has not been estimated for multiple age groups and it may differ between younger and older adults), we restricted our study to adults in their early to mid-life (25-64 years) at baseline in line with the original study that estimated effect of takeaway exposure on BMI?^33^.

Our modelling assumed that during the course of the study, the BMI of adults aged 65 plus were no longer influenced by a change in exposure to takeaways, with any differences in BMI between business-as-usual and the intervention scenario remaining constant in this cohort after this point. This decision was informed by recent literature on dietary intake that showed how in the UK, younger adults (aged 19-29 years) were five times as likely to eat takeaway meals at home relative to adults aged 70 years+^37^ . Thus, we took a cautious approach to ensure we did not overestimate any potential impact of reduction in exposure to takeaway outlets on BMI. Disease incidence estimates were based on time lags from the effect of BMI changes from takeaway exclusion zones. A time lag of five years was assigned for diabetes and cardiovascular diseases based on WHO estimates of reversal of stroke and heart disease^38^, 10 years for cancer based on cohort study findings examining intentional weight loss and breast cancer risk ^39^, and one year for all other diseases. A schematic diagram of our analytical strategy is shown in figure 1.

**Figure 1:**
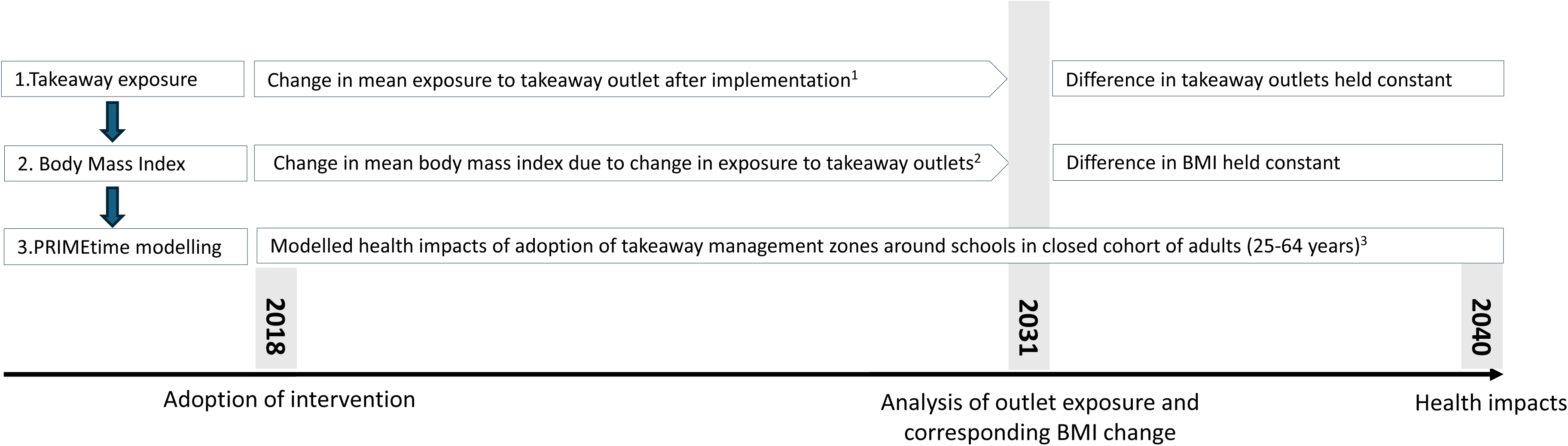
Schematic diagram of analysis strategy. ^1^Change in mean exposure to takeaways (by 2031) is calculated by comparing the difference in outlet exposure from a business-as-usual model (see Liu et al, 2024) to an intervention that reduces outlet growth between by 50%. ^2^ For each additional takeaway an individual is exposed to, mean BMI increases by 0.0241 kg/m^2.^ See Burgoine et al, 2014 ^3^For PRIMEtime modelling, the oldest age of a cohort member would be aged 64 years old at baseline (2018) and who would be 86 years old by 2040. Some adults will be lost to follow-up, for example due to premature mortality.

In sensitivity analyses, relative to business-as-usual growth, we modelled impacts of policy adoption under a “perfect implementation” scenario where there was a total (100%) reduction in new takeaway growth in the takeaway management zones.

## Results

Demographic characteristics of the six LAs are described in table 1. Wandsworth, a major urban LA in London, has a population density approximately 50 times higher than Fenland, an LA that is mainly rural. Urban LAs had populations with a higher proportion of younger adults (aged 25-44 years). For example, 69% of the population of Wandsworth is within this age group, whereas it constitutes only 46% of Fenland’s population.

### Takeaway exposure following adoption of management zones

Mean exposure to takeaways at baseline varied between the six LAs, with populations of rural LAs (North Somerset and Fenland) exposed on average to approximately two-thirds fewer takeaways than in other more urban LAs (Table 2). Adoption of takeaway management zones, assuming realistic implementation, led to exposure to fewer takeaways on average, per person, across all LAs relative to business as usual, with the highest absolute reductions in more urban areas. For example, in Manchester, realistic implementation was estimated to reduce average exposure to 28.4 (95% CI 25.8, 31.0) fewer new takeaways per person by 2031, relative to business as usual. Reductions in takeaway exposure were lower in other LAs, with exposure to 3.2 (95% CI 1.98, 4.43) fewer new takeaways in Fenland, relative to business as usual. Reductions were stronger under an optimistic implementation scenario, and strongest under perfect implementation, where we estimated in Manchester that takeaway management zones with those stringencies would lead to exposure to 42.6 (95% CI 38.7, 46.5) and 56.8 (95% CI 51.6, 61.9) fewer takeaways per person, relative to business as usual (Table S1).

**Table 2:** Estimated difference in mean number of takeaways an adult is exposed to in 2040 due to the intervention compared to business-as-usual.

| Local authority | Baseline exposure in 2018<sup>4</sup> | Mean difference in outlet exposure per person compared to business-as-usual scenario |
| --- | --- | --- |
| Wandsworth | 73.5 | -12.6(-9.51, -15.6) |
| Manchester | 91.4 | -28.4(-25.8, -31.0) |
| Sheffield | 74.9 | -21.4(-17.3, -25.5) |
| Blackburn with Darwen | 66.6 | -15.7(-9.43, -22.0) |
| North Somerset | 18.6 | -4.09(-3.50, -4.69) |
| Fenland | 17.7 | -3.20(-1.98, -4.43) |

<sup>1</sup>Upper and Lower confidence intervals are indicated in brackets.
<sup>2</sup>Trajectories of takeaway growth were assumed to increase until 2031 and then stabilise between 2031-2040
<sup>3</sup>The intervention was based on a realistic scenario where new takeaway growth reduces by 50% each year following the intervention.
<sup>4</sup> Estimated Outlet exposure (from home, work and commuting) in 2018

### Changes in mean BMI after takeaway management zone implementation

Realistic implementation was associated with an estimated per person reduction in BMI that was greatest in Manchester (0.68kg/m^2^; 95% CI 0.62, 0.75) and lowest in Fenland (0.08kg/m^2^; 95% CI 0.05, 0.11) in 2031 compared to business-as-usual, and that was overall greater in more urban LAs (Table 3). These patterns were consistent, but effects were stronger under an optimistic implementation scenario, and stronger still under perfect implementation, where in Manchester the intervention would result in BMI of 1.03 kg/m^2^ (95%CI 0.93, 1.25) and 1.37 kg/m^2^ (95%CI 1.24, 1.49) lower respectively, relative to business as usual (Table S2). Across the spectrum of deprivation within each LA, mean population BMI change appeared stable, but with some indication that in rural areas, greater reductions in BMI may occur in the least deprived areas (Table S3).

**Table 3:** Change in mean BMI in the adult population (2018 to 2040) in six specified local authorities, following implementation of takeaway management zones in 2018.

| Local authority | Baseline Obesity level (%)<sup>3</sup> | Estimated change in BMI (kg/m<sup>2</sup>) |
| --- | --- | --- |
| Wandsworth | 14.4 | -0.30 (-0.23, -0.38) |
| Manchester | 25.4 | -0.68 (-0.62, -0.75) |
| Sheffield | 25.3 | -0.52(-0.42, -0.61) |
| Blackburn with Darwen | 23.0 | -0.38(-0.23, -0.53) |
| North Somerset | 23.0 | -0.10(-0.08, -0.11) |
| Fenland | 40.1 | -0.08(-0.05, -0.11) |

<sup>1</sup> Trajectories of BMI were assumed to change until 2031 and then stabilise between 2031-2040
<sup>2</sup> The intervention was based on a realistic scenario where new takeaway growth reduces by 50% each year following the intervention.
<sup>3</sup> Percentage of adults aged 18 + who are living with obesity

### Change in prevalence of obesity, QALYs and healthcare cost savings to 2040

We estimated reductions in obesity prevalence for all LAs, compared to business as usual. In males, percentage point (PP) reductions in obesity prevalence ranged from 2.3PP (95% UI 2.9, 1.7) in Manchester to 1.5PP (95% UI 1.9, 1.1) in Wandsworth (Table 4). In females these reductions ranged from 1.9PP (95% UI 2.4, 1.4) in Manchester and Sheffield to 1.5PP (95% UI 10.9, 1.2) in Wandsworth. Our models also estimated gains in total QALYs for all LAs, which ranged from a gain of 249 QALYs per 100,000 population for adults living in Manchester, to a gain of 194 QALYS per 100,00 adults living in North Somerset. In terms of healthcare cost savings, these ranged from £2.02 million saved per 100,000 adults in Manchester to £1.65 million saved per 100,000 adults living in Fenland over the 22 year period. In sensitivity analysis, healthcare cost savings, QALYs and change in prevalence of obesity were all approximately twice that observed under a realistic implementation scenario in the main analysis (Table S4).

**Table 4:**
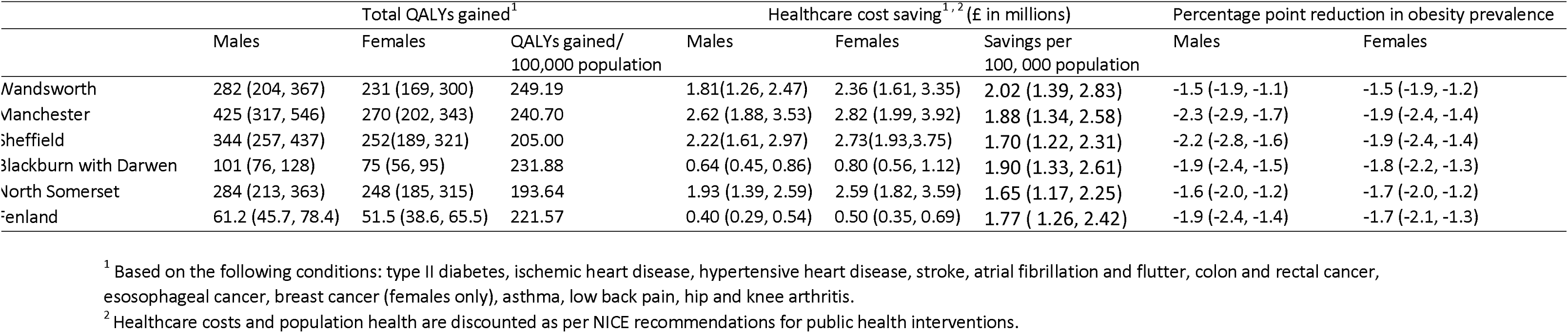
Impact of the intervention on quality adjusted life years (QALYs), healthcare cost savings and obesity prevalence in the adult population (2018 to 2040) in six specified local authorities.

### Change in incident cases of disease to 2040

The largest estimated reductions in cases of disease were for type II diabetes, with an estimated reduction of 1013 (95% UI 1285, 735) male and 837 (95% UI 1048, 634) female cases per 100,000 population, by 2040, in Blackburn with Darwen (Table 5). Reductions in all forms of cardiovascular disease were also observed, with reductions in IHD (e.g. Blackburn with Darwen, males: 153 cases/100,000 population, 95% UI 117, 192) and atrial fibrillation (e.g. Blackburn with Darwen, males: 73 cases/100,000 population, 95% UI 48, 102) strongest in all LAs, and consistently more pronounced in males. Improvements for respiratory health, with marked reductions in asthma, particularly for females (e.g. Blackburn with Darwen: 402 cases/100,000 population, 95% UI 220, 603), were also observed. Smaller reductions were estimated for oesophageal, breast, and colon and rectum cancers across all LAs. Of all cancers, case reductions were greatest for breast cancer. In terms of impacts on musculoskeletal disease, reductions were estimated for low back pain and more so for females than males (e.g. Blackburn with Darwen: 326 cases/100,000 population, 95% UI 17, 644). Small increases in incidence rates for hip and knee osteoarthritis were consistently estimated for both sexes in all LAs. In sensitivity analysis, perfect implementation (i.e. no new takeaways being allowed to open after policy adoption) resulted in an almost doubling of reductions, in disease incidence across all LAs, relative to realistic implementation (Table S5).

**Table 5:** Change in incident cases of disease /100,000 adult population (2018 to 2040), in six specified local authorities.

|  | Blackburn with Darwen | Fenland | Manchester | Sheffield | North Somerset | Wandsworth |
| --- | --- | --- | --- | --- | --- | --- |
| **Males** |  |  |  |  |  |  |
| **Metabolic** |  |  |  |  |  |  |
| Type II diabetes | -1013 (-1285, -753) | -995 (-1262, -740) | -964 (-1565, -870) | -803(-1023, -594) | -804 (-1018, -600) | -1206 (-1565, -870) |
| **Cardiovascular disease** |  |  |  |  |  |  |
| Ischaemic heart disease | -153 (-192, -117) | -105(-131, -80.6) | -124 (-157, -94.1) | -118 (-149, -90.0) | -91.1 (-114, -70.3) | -99.2 (-125, -75.3) |
| Hypertensive heart disease | -8.16 (-13.5, -3.22) | -8.32 (-13.6, -3.45) | -6.89 (-11.9, -2.13) | -7.91 (-13.1, -3.09) | -8.42 (-13.7, -3.59) | -6.73 (-11.9, -2.03) |
| Stroke | -9.32 (-12.5, -6.41) | -15.9 (-21.1, -11.0) | -21.1 (-28.3, -14.5) | -17.3 (-23.3, -11.9) | -15.2 (-20.1, -10.5) | -18.9 (-25.3, -12.9) |
| Atrial fibrillation & flutter | -72.5 (-102, -47.8) | -61.7 (-86.6, -40.7) | -60.6 (-85.5, -39.8) | -59.7 (-83.8, -39.4) | -57.3 (-80.3, -4.17) | 62.0 (-87.7 -40.6) |
| **Cancer** |  |  |  |  |  |  |
| Colon & rectum Cancer | -1.05 (-1.57, -0.52) | -1.58 (-2.77, -0.79) | -1.59 (-2.41, -0.83) | -1.76 (-2.64, -0.93) | -1.22 (-1.79, -0.69) | -0.91. (-1.31, -0.50) |
| Oesophageal | -0.03 (-0.05, -0.03) | -3.16 (-4.35, -1.98) | -3.28 (-4.54, -2.24) | -2.94 (-4.04, -2.02) | -3.58 (-5.00, -2.44) | < 0.01 (0.01, 0.01) |
| **Respiratory** |  |  |  |  |  |  |
| Asthma | -196 (-293, -107) | -187 (-283, -101) | -192 (-288, -105) | -169 (-252, -93.4) | -182 (-278, -97.2) | -238 (-366, -125) |
| **Musculo-skeletal** |  |  |  |  |  |  |
| Low back pain | -272 (-532, -21.0) | -278 (-556, -5.03) | -332 (-650, -29.0) | -277 (-534, -31.6) | -332 (-676, 1.58) | -249 (-534, 22.2) |
| Hip osteoarthritis | 0.52 (0.52, 0.79) | 0.40 (0.40, -0.40) | 0.6 (0.47, 0.73) | 0.55 (0.41, 0.62) | 0.15 (0.15, 0.22) | 0.30 (0.20, -0.81) |
| Knee osteoarthritis | 2.62 (2.10, 2.62) | 1.98 (1.58, 2.37) | 2.74 (2.20, 3.34) | 2.41 (1.92, 2.96) | 0.97 (0.75, 1.19) | 1.21 (0.91, 1.51) |
| **Females** |  |  |  |  |  |  |
| **Metabolic** |  |  |  |  |  |  |
| Type II diabetes | -837 (-1048, -634) | -987 (-1228, -754) | -725(-911, -546) | -677(-847, -513) | -801 (-997, -612) | -879(-1116, -648) |
| **Cardiovascular disease** |  |  |  |  |  |  |
| Ischaemic heart disease | -49.8 (-62.3, -38.3) | -39.2 (-48.9, -30.2) | -41.8 (-52.7, -31.8) | -41.1 (-51.7, -31.4) | -34.4 (-42.8, -26.6) | -31.4 (-39.7, -23.9) |
| Hypertensive heart disease | -5.47 (-8.72, -2.38) | -5.92 (-9.18, -2.77) | -4.68 (-7.52, -1.86) | -5.37 (-8.40, -2.34) | -6.18 (-9.73, -2.91) | -4.40 (-7.40, -1.59) |
| Stroke | -19.0 (-25.6, -13.5) | -15.2 (-20.4, -10.8) | -20.1 (-27.2, -13.9) | -16.7 (-22.5, -11.6) | -15.3 (-20.4, -10.9) | -15.2 (-20.7, -10.5) |
| Atrial fibrillation & flutter | -32.7 (-45.9, -21.6) | -30.9 (-43.3, -20.5) | -30.9 (-43.3, -20.5) | -27.2 (-38.1, -18.0) | -31.8 (-44.6, -21.1) | -24.1 (-34.1, -15.8) |
| **Cancer** |  |  |  |  |  |  |
| Colon & rectum Cancer | -1.02(-1.50, -0.57) | -0.78 (-1.56, -0.39) | -0.95 (-1.40, -0.53) | -0.96 (-1.45, -0.55) | -1.14(-1.71, -0.64) | -0.67(-1.03, -0.37) |
| Oesophageal | <0.01 (0.01, 0.01) | -0.78 (-0.78, -0.39) | <0.01 (0.01, 0.01) | -0.48 (-0.69, -0.34) | <0.01 (0.01, 0.01) | <0.01 (0.01, 0.01) |
| Breast Cancer | -6.43 (-8.63, -4.49) | -6.65 (-8.99, -4.69) | -6.56 (-8.82, -4.58) | -6.61 (-8.88, -4.61) | -6.68 (-8.95, -4.69) | -6.28 (-8.43, -4.40) |
| **Respiratory** |  |  |  |  |  |  |
| Asthma | -402 (-603, -220) | -327 (-490, -179) | -361 (-542, -199) | -325 (-485, -180) | -318 (-483, -171) | -444 (-681, -235) |
| **Musculo-skeletal** |  |  |  |  |  |  |
| Low back pain | -326 (-644, 16.5) | -312 (-613, 16.7) | -325 (-638, -23.8) | -319 (-618, 31.2) | -316 (-613, 1.51) | -318 (-666, 11.3) |
| Hip osteoarthritis | 0.26 (0.26, 0.26) | 004 (0.03, 0.05) | 0.22 (0.14, 0.22) | 0.14 (0.14, 0.21) | 0.14 (0.14, 0.21) | 0.09 (0.09, 0.09) |
| Knee osteoarthritis | 1.06 (0.79, 1.32) | 0.78 (0.78, 1.17) | 1.08 (0.86, 1.37) | 0.96 (0.76, 1.17) | 0.78 (0.78, 1.17) | 0.47 (0.37, 0.66) |

## Discussion

### Summary of findings

Our findings suggest that takeaway management zones around schools could make a substantive contribution to improving adult health and associated healthcare costs. We estimated that this intervention would reduce prevalence of obesity by 1.5 to 2.3 percentage points by 2031, leading to improvements in BMI-related health outcomes to 2040. These estimates were forecast to result in reductions in incidence of a range of diseases by 2040, including type II diabetes, cardiovascular diseases, and asthma. Estimated healthcare cost savings and gains in QALYs were similar in magnitude across LAs, with healthcare savings ranging between £1.65 – 2.02 million per 100,000 population and gains in QALYs ranging from between 194 to 241 QALYs gained/ 100,000 population in North Somerset and Wandsworth, respectively. We also found that more stringent implementation of the policy, in alternate optimised or perfect scenarios, would result in even greater population health benefits.

### Comparison with other studies

This is the first study attempting to estimate health impacts of takeaway exclusion zones, making it challenging to make direct comparisons with other studies. However, reductions in obesity prevalence in relation to takeaway management zones were consistent across LAs and in line with a number of other studies that have found a relationship between higher exposure to takeaways and increased BMI or risk of obesity in adults^11–13^. Meaningful reductions were estimated for future incidence of 12 obesity-related diseases to 2040 across all LAs irrespective of rural-urban classification. The most pronounced reductions, in all LAs, were in incidence of type II diabetes, which in males ranged from reductions of 803 cases/100,000 population in largely rural North Somerset to 1206 cases/ 100,000 population in urban Wandsworth. Consistent with this finding, previous studies have shown a positive association between residential takeaway exposure and prevalence of type II diabetes^7,9^. This is an important finding because aside from older age, type II diabetes incurs the biggest financial cost of any single disease to NHS healthcare, accounting for 8% of secondary care costs and occupying 17% of hospital day-beds^40^. Our estimates also showed substantial reductions in incidence of cardiovascular diseases in response to adoption of management zones. The largest reductions in incident cases were seen in ischaemic heart disease (IHD), with smaller reductions in stroke and hypertensive heart disease. Consistent with this finding, a recent systematic review highlighted evidence of a relationship between takeaway exposure and cardiovascular disease risk^41^. Furthermore, another study found that incidence of CVD and to a lesser extent stroke, was also higher in adults exposed to more takeaways, which mirrors our observations^8^. Our model also estimated meaningful reductions in the incidence of some cancers, asthma, and low back pain. While research on the link between takeaway exposure and these conditions is lacking, each has been found to be associated with living with obesity^42–44^. We found no significant differences between adoption of the intervention and BMI change by level of deprivation within LAs. This is similar to a previous study that found no differential impact of takeaway exposure across levels of household income^13^.

### Interpretation of findings

Recent data from the Health Survey for England suggested that approximately 26% of adults are obese, with the highest prevalence in age-groups 45-74 years.^45^ This suggests that adults in this age group may be an important group to target, especially given the relationship between obesity and disability and chronic disease in older adults^46^. However, while the significant reductions in obesity prevalence estimated by our models are encouraging (e.g. 1.9 PP in females in Sheffield), they also illustrate the need for a broader set of diet-related interventions to further reduce prevalence of obesity. Many public health interventions are cost saving ^47^ and while the financial costs of the implementation of takeaway management zones were not included in our study and should be integrated into future analyses, healthcare savings were estimated to range from £1.65 million per 100,000 population in North Somerset to £2.02 million per 100,000 population in Wandsworth by 2040. If sustained over a period of 22 years, our modelling also showed that takeaway management zones could add between 101 (Blackburn) and 425 (Manchester) QALYs for males alone, suggesting that the intervention has the potential to make meaningful improvements to the quality of life of whole populations. Our models also estimated slight increases in incidence of knee and hip osteoarthritis. While BMI is associated with osteoarthritis^48^, this finding can be explained by the higher proportion of older adults surviving in the population because of the intervention^49^. While our findings estimate larger BMI reductions in more urban LAs, our modelling of health impacts does not mirror this difference between urban and rural areas with incidence of non-communicable diseases and change in obesity prevalence, healthcare savings and QALYs per person. This finding may reflect differences in the demographics of selected LAs including baseline obesity levels, deprivation and age which are risk factors for poor health.

### Study limitations

#### Limitations: Forecasting model of takeaway growth

Our study makes use of unique forecasts of long-term population exposure to takeaways in the absence of intervention, in six different types of LAs, based on continuation of pre-existing trends in takeaway growth. As the intervention can only stop future growth, the benefits of the intervention are contingent on continued growth (in the absence of intervention), and this is inherently uncertain. For example, to what extent will growth in numbers of physical premises continue (and to be important) if online takeaway delivery use continues to rise. Further detail on limitations of this method have been published previously^32^. There is also uncertainty around the effectiveness of the implementation. To address this, in addition to a core scenario based on recent estimates of real-world impact^50^, we also provided estimates based on alternative scenarios.

#### Limitations: generalisability

Our findings are not readily generalisable to children. In this study we focussed on the adult population, in part because previously published associations between takeaway exposure and BMI were based on similar working-age UK adults^11^. Also because takeaway management zones affect a wide geographical area, it is reasonable to assume they will also impact the adult population. Moreover, the geographic and social determinants of takeaway consumption in children may be different and this should be the subject of future research. While observational studies in children show an association between takeaway consumption and energy intake, no corresponding association between takeaway consumption and body weight has been observed, perhaps because energy demands tend to be higher for growth and development^51,52,53^Evidence on the relationship between exposure to takeaways and body weight in older populations is also currently lacking, thus our models did not include adults who were aged 65 years and over at study baseline. However, a study using data from the UK National Diet and Nutrition Survey found that adults aged 70 years and over were one fifth as likely to eat takeaway meals at home compared to young adults aged 19-29 years. This supports the idea that dietary behaviours are subject to change over the lifecourse^37^, necessitating further modelling of intervention impacts in this older age group.

#### Limitations: PRIMEtime modelling

The PRIMEtime model excludes some important diseases associated with BMI, including depression and dementia, potentially leading to our results being an underestimation of effect sizes for savings in healthcare costs and QALYs^52,53^. In choosing to model a closed cohort we will have potentially further underestimated the health and heathcare cost savings. BMI is also positively linked to need for social care provision^54^ however we have not modelled social care costs. In the UK, social care costs (in contrast to healthcare costs) are borne by the local authority, and so the returns to the body that bears the risks and costs of the intervention are not quantified here.

#### Limitations: Online food delivery

We were unable to account for the impact of online food delivery services (e.g. Just Eat, Deliveroo), which may attenuate the relationship between takeaway management zones and health. These fast-growing services are likely to increase the availability of takeaway food, which the intervention was designed to reduce, thereby reducing its impact. In one UK study, online food delivery services were used at least once per week by approximately 15% of adults in 2018^55^ and there is evidence that access is unequal between urban and rural areas^56^From 2020 to 2022, access to online delivery takeaways was found to have increased by 10% for those living in the most deprived areas of England^57^. Adults living in the UK who have access to the greatest number of takeaways online were also found to have the greatest odds of using online food delivery services^58^ ^55^. Future research should consider the possibility that place-based interventions such as management zones may to some extent be undermined by new modes of takeaway food purchasing.

### Policy implications and future directions

A lack of evidence of health benefits associated with the adoption of takeaway management zones around schools has been cited as a barrier to policy adoption and effective implementation^27,28^. Building on recent studies that have observed the retail impacts of policy adoption^26,59^, our modelling work now provides evidence on the population health impacts that could be achieved through the adoption and (even imperfect) implementation of takeaway management zones around schools. We also showed how stricter, perfect or even optimised implementation (preventing takeaway growth by 100% and 75%, respectively) would result in even greater benefits to 2040. Local decision makers should therefore remain diligent in the strict implementation of takeaway management zones if optimum population health is to be achieved.

In addition to a range of health benefits we also modelled economic benefits associated with the adoption of takeaway management zones around schools, which were achieved through a reduction in healthcare costs. Although these economic benefits may not accrue locally, these cost savings are important evidence for those working in LAs who seek to understand the wider health benefits of management zones^60^. It is still possible however, as argued by inspectors from the national planning inspectorate, that management zones could be detrimental to the economy, through denying business growth and curtailing employment opportunities^61^. As public health interventions are liable to legal challenge under the principle of proportionality,future work should include a full economic analysis, considering both health and social care costs and benefits, alongside these other economic considerations. Future studies should also account for the continued emergence and growth of online food delivery platforms, which could diminish the health impacts of this intervention.

### Conclusions

The is the first study to model the health impacts of the adoption and implementation of takeaway management zones (sometimes referred to by LAs as “exclusion zones”) around schools in . In response to a realistic intervention scenario and across a range of different types of LAs, we found meaningful reductions in population-level BMI and obesity prevalence, and in a variety of associated non-communicable diseases including incidence of type II diabetes, cardiovascular diseases, cancers, asthma and low back pain, to the year 2040. We also found associated health-related benefits including gains in QALYs and savings in healthcare costs. Takeaway management zones around schools may be an effective population-level intervention to improve diet-related health in adults in the UK ^62^

## Funding and acknowledgements

This study is funded by the National Institute for Health Research (NIHR) Public Health Research Programme (Project number: NIHR130597). The views expressed are those of the author(s) and not necessarily those of the NIHR or the Department of Health and Social Care. JR, MK, BL, AS, SJS, MW, NR, JA and TB were supported by the Medical Research Council (grant number MC_UU_00006/7). OM is supported by a UKRI Future Leaders Fellowship (MR/T041226/1)). For the purpose of open access, the author has applied a Creative Commons Attribution (CC BY) licence to any Author Accepted Manuscript version arising.

## Data Availability

Derived data on projection of takeaway outlet growth is available upon request.

## Notes

### Competing Interest Statement

The authors have declared no competing interest.

