## Supplemental Tables for "Health impacts of takeaway management zones around schools in six different local authorities across England: a public health modelling study using PRIMEtime"

**Supplementary Tables**

Table S1. Estimated difference in mean number of takeaways a person is exposed to due to the intervention compared to business as usual.

Table S2. Change in mean BMI for the adults aged 25-64 years across 6 specified local authorities in England, by 2031, following implementation of takeaway exclusion zones in 2018.

Table S3. Estimated changes in mean population BMI across the six specified local authorities in England, overall and by deprivation quintile with LA, following implementation of a takeaway exclusion zone assuming 100% stringency.

Table S4. Impact of the intervention, on QALYs, health care costs and change in obesity prevalence in the adult population from 2018-2040 in specified local authorities.

Table S5. Change in incident cases of disease per 100,000 adult population (2018 to 2040), in specified local authorities as a result of the intervention, assuming 100% stringency

Table S1: Estimated difference^1^ in mean number of takeaways a person is exposed to due to the intervention^2^ compared to business as usual.

|  | Baseline  Exposure in 2018^3^ | Estimated difference in outlet exposure/person in 2031 compared to business as usual scenario | |
| --- | --- | --- | --- |
|  | | Optimistic^1^ | Perfect^1^ |
| Wandsworth | 73.5 | -18.8(-14.3, -23.4) | -25.1(-19.0, -31.2) |
| Manchester | 91.4 | -42.6(-38.7, 46.5) | -56.8(-51.6, -61.9) |
| Sheffield | 74.9 | -32.1(-26.0, -38.2) | -42.8(-34.7, -51.0) |
| Blackburn with Darwen | 66.6 | -23.6(-14.1, -33.0) | -31.4(-18.9, -44.0) |
| North Somerset | 18.6 | -6.14(-5.24, -7.04) | -8.19(-6.99, -9.38) |
| Fenland | 17.7 | -4.80(-2.96, 6.64) | -6.40(-4.00, -8.85) |

^1^Upper and Lower confidence intervals are indicated in brackets.

^2^The intervention here is based on an optimistic and perfect scenario where new takeaway growth reduces by 75% and 100% each year, respectively, following the intervention.

^3^Estimated Outlet exposure (from home, work and commuting) in 2018

|  |  | Estimated changes in BMI (kg/m^2^) | |
| --- | --- | --- | --- |
|  | Baseline Obesity levels (%)^2^ | Optimistic^3^ | Perfect^3^ |
| Wandsworth | 14.4 | -0.45(-0.34, -0.56) | -0.61(-0.46, -0.75) |
| Manchester | 25.4 | -1.03(-0.93, -1.25) | -1.37(-1.24, -1.49) |
| Sheffield | 25.3 | -0.77(-0.63, -0.92) | -1.03(-0.84, -1.23) |
| Blackburn with Darwen | 23.0 | -0.57(-0.34, -0.79) | -0.76(-0.45, -1.06) |
| North Somerset | 23.0 | -0.15(-0.13, -0.17) | -0.20(-0.17, -0.23) |
| Fenland | 40.1 | -0.12(-0.07, -0.16) | -0.15(-0.10, -0.21) |

Table S2. Change in mean BMI for the adults aged 25-64 years across 6 specified local authorities in England, by 2040^1^, following implementation of takeaway exclusion zones in 2018.

^1^ Trajectories of takeaway growth were assumed to increase until 2031 and then stabilise from 2031-2040.

^2^Percentage of adults aged 18 + who are living with obesity.

^3^ The intervention here is based on an optimistic and perfect scenario where new takeaway growth reduces by 75% and 100% each year, respectively, following the intervention.

Table S3. Estimated changes in mean population BMI^1^ in a closed cohort of adults aged 25-64 (2018-2031) across the six specified local authorities in England, overall and by deprivation quintile with LA, following implementation of a takeaway exclusion zone assuming 100% stringency.

| Local authority | Manchester | Wandsworth | Sheffield | North Somerset | Fenland | Blackburn with Darwen |
| --- | --- | --- | --- | --- | --- | --- |
| Overall | -1.37(-1.24, -1.49) | -0.69(-0.52, -0.87) | -1.10(-0.88, -1.32) | -0.21(-0.18, -0.24) | -0.17(-0.10, -0.23) | -0.81(-0.48, -1.14) |
| Quintile 1 (least deprived) | -1.41(-1.29, -1.53) | -0.67(-0.45, -0.88) | -1.01(-0.87, -1.15) | -0.60(-0.51, -0.70) | -0.32(-0.19, -0.45) | -1.06(-0.65, -1.47) |
| Quintile 2 | -1.63 (-1.48, -1.79) | -0.67(-0.50, -0.85) | -1.24(-1.04, -1.44) | -0.49(-0.41, -0.57) | -0.18(-0.12, -0.24) | -1.00(-0.64, -1.36) |
| Quintile 3 | -1.34 (-1.23, -1.45) | -0.72(-0.59, -0.86) | -1.29 (-1.04, -1.54) | -0.27(-0.24, -0.30) | -0.14(-0.10, -0.19) | -0.90(-0.57, -1.24) |
| Quintile 4 | -1.47(-1.36, -1.58) | -0.70(-0.49, -0.90) | -1.33 (-1.06, -1.60) | -0.18(-0.15, -0.22) | -0.11(-0.07, -0.15) | -1.33(-1.13, -1.53) |
| Quintile 5 (most deprived) | -1.56 (-1.14, -1.98) | -0.67(-0.49, -0.85) | -1.21(-0.92, -1.51) | -0.12(0.17, -0.41) | No Q5 in Fenland | -0.58(-0.29, -0.88) |

Table S4: Impact of the intervention, on QALYs, health care costs and change in obesity prevalence in the adult population from 2018-2040 in specified local authorities.

|  | Total QALYs | | Healthcare cost savings^1^ (£ in millions) | | Change in prevalence of obesity (PP) | |
| --- | --- | --- | --- | --- | --- | --- |
|  | Males | Females | Males | Females | Males | Females |
| Optimistic (75% stringency) | | | | | | |
| Wandsworth | 420 (309, 551) | 344 (253, 447) | 2.68(1.86, 3.71) | 3.49 (2.34, 4.93) | -2.2 (-1.7, -2.8) | -2.4 (-1.9, -3.0) |
| Manchester | 635 (478, 812) | 403 (302, 512) | 3.89 (2.75, 5.21) | 4.19 (2.89, 5.80) | -3.3 (-4.2, -2.5) | -2.9 (-3.7, -2.2) |
| Sheffield | 514 (387, 654) | 377(284, 480) | 3.31(4.41, 2.34) | 4.05(2.82,5.60) | -3.3 (-4.1, -2.5) | -2.9 (-3.7, -2.2) |
| Blackburn with Darwen | 151 (113, 192) | 111 (84, 143) | 0.95 (0.67, 1.27) | 1.2 (0.82, 1.66) | -2.9 (-3.7, -2.2) | -2.8 (-3.5, -2.1) |
| North Somerset | 424 (321, 542) | 370 (279, 470) | 2.86 (2.03, 3.82) | 3.84 (2.67, 5.31) | -2.3(-2.8, -1.8) | -2.5 (-3.1, -1.9) |
| Fenland | 91.5 (69.2, 117) | 77.0 (58.2, 97.5) | 0.59 (0.42,0.80) | 0.74 (0.52, 1.02) | -2.8 (-3.5, -2.2) | -2.7 (-2.1, -3.4) |
| Perfect (100% stringency) | | | | | | |
| Wandsworth | 550 (392, 733) | 450 (325, 592) | 3.50 (2.37, 4.96) | 4.56 (2.97, 6.53) | -2.9 (-2.3, -3.6) | -3.2 (-2.5, -4.0) |
| Manchester | 831 (609, 1077) | 527 (391, 674) | 5.09 (3.54, 7.07) | 5.49 (3.67, 7.68) | -4.4 (-5.4, -3.4) | -3.8 (-4.8, -3.0) |
| Sheffield | 673(502, 868) | 494 (368, 633) | 4.34 (3.04, 5.98) | 5.31 (3.58, 7.41) | -4.3 (-3.3, -5.3) | -3.9 (-3.0, -4.8) |
| Blackburn with Darwen | 198 (147, 256) | 146 (109, 188) | 1.24 (1.73, 0.86) | 1.56(2.19, 10.4) | -3.9 (-4.8, -3.0) | -3.6 (-4.5, -2.8) |
| North Somerset | 555 (412, 717) | 485 (361, 625) | 3.74 (2.62, 5.21) | 5.03 (3.37, 7.00) | -3.0(-3.7, -2.3) | -3.3(-4.0, -2.5) |
| Fenland | 120 (88, 155) | 120 (155, 88) | 0.78 (10.8, 0.55) | 0.78 (10.8, 0.55) | -3.7 (-4.4, -2.8) | -3.6 (-4.4, -2.8) |

|  | Blackburn with Darwen | Fenland | Manchester | Sheffield | North Somerset | Wandsworth |
| --- | --- | --- | --- | --- | --- | --- |
| **Males** | | | | | | |
| **Metabolic** |  |  |  |  |  |  |
| Type II diabetes | -1961 (-2503, -1452) | -1926 (-2453, -1428) | -1864 (-2396, -1354) | -1553(-1982, -1143) | -1556 (-1979, -1158) | -2327 (-3054, -1657) |
| **Cardiovascular disease** |  |  |  |  |  |  |
| Ischaemic heart disease | -301 (-373, -233) | -206 (-254, -159) | -244 (-306, -187) | -231 (-289, -179) | -179 (-220, -139) | -194 (-242, -142) |
| Hypertensive heart disease | -16.0 (-26.5, -5.77) | -16.2 (-26.9, -6.33) | -13.4 (23.6, -3.94) | -15.4 (-25.8, -5.50) | -16.4 (-27.1, -6.64) | -13.0 (-23.6, -3.83) |
| Stroke | -18.1 (-24.7, -12.9) | -31.3 (-41.9, -22.2) | -41.1 (-56.0, -28.7) | -33.8 (-45.9, -23.8) | -29.6 (-39.7, -21.1) | -36.6 (-172, -25.5) |
| Atrial fibrillation & flutter  **Cancer** | -142 (-201, -93.9) | -121 (-170, -80.3) | -119 (-168, -78.2) | -117 (-165, -77.7) | -112 (-158, -74.3) | 121 (-172, -79.5) |
| Colon & rectum Cancer  Esophageal  **Respiratory** | -2.10 (-2.89, -1.05)  -0.05 (-0.07, -0.04) | -3.56 (-5.14, -1,98)  -5.93 (-8.31, -3.96) | -3.14 (-4.68, -1.60)  -6.35 (-8.89, -4.21) | -3.44 (-5.16, -1.86)  -5.70 (-7.90, -3.85) | -3.58 (-5.30, -1.94)  -6.94 (-9.78, -4.55) | -1.71 (-2.52, -0.91)  < 0.01 (0.01, 0.01) |
| Asthma  **Musculo-skeletal** | -380 (-579, -205) | -363 (-558, -193) | -373 (-569, -200) | -329 (-500, -178) | -352 (-543, -184) | -458 (-713, -236) |
| Low back pain | -533 (-1044, -10.8) | -542 (-1096, -17.0) | -651 (-1270, -19.4) | -544 (-1045, -30.0) | -502 (-1045, 50.7) | -484 (-1033, 64.3) |
| Hip osteoarthritis  Knee osteoarthritis | 1.05 (1.05, 1.31)  5.51 (4.20, 6.56) | 0.79 (0.79, -1.19)  3.96 (3.16, 4.75) | 1.20 (0.94, 1.47)  5.34 (4.21, 6.55) | 1.03 (0.82, 1.24)  4.74 (3.71, 5.77) | 0.82 (0.67, 0.97)  3.88 (2.99, 4.70) | 0.50 (0.40, 0.61)  2.42 (1.82, 3.03) |
| **Females** | | | | | | |
| **Metabolic** |  |  |  |  |  |  |
| Type II diabetes | -1622 (-2049, -1235) | -1915 (-2400, -1474) | -1405 (-1778, -1059) | -1314 (-1657, -1000) | -1555 (-1952, -1199) | -1700 (-2174, -1245) |
| **Cardiovascular disease** |  |  |  |  |  |  |
| Ischaemic heart disease | -97.6 (-122, -75.7) | -77.0 (-95.8, -60.2) | -82.0 (-103, -63.3) | -80.6 (-101, -62.5) | -67.4 (-83.8, -52.7) | -61.5 (-77.3, -47.5) |
| Stroke | -37.3 (-50.3, -26.5) | -29.7 (-40.3, -21.1) | -39.1 (-53.9, -27.0) | -32.6 (-44.5, -22.7) | -29.8 (40.1, -21.5) | -29.5 (-40.6, -20.3) |
| Hypertensive heart disease | -10.6 (-17.2, -4.50) | -11.7 (-18.0, -5.08) | -9.13 (-14.7, -3.31) | -10.5 (-16.6, -4.34) | -12.1 (-19.0, -5.54) | -8.53 (-14.4, -2.91) |
| Atrial fibrillation & flutter  **Cancer** | -64.0 (-90.5, -42.3) | -60.6 (-85.2, -40.3) | -54.9 (-77.7, -36.2) | -53.4 (-75.3, -35.3) | -62.4 (-88.1 -41.4) | -47.1 (-66.9, -31.0) |
| Colon & rectum Cancer  Esophageal  Breast Cancer  **Respiratory** | -2.12 (-2.91, -1.06)  <0.01 (0.01, 0.01)  -12.7 (-16.9, -8.73) | -1.95 (-2.74, -1.17)  -1.17 (-1.95, -0.78)  -12.9 (-17.2, -8.99) | -1.87(-2.73, -1.08)  <0.01 (0.01, 0.01)  -12.9 (-17.2, -8.99) | -1.93 (-2.89, -1.10)  -1.03 (-1.38, -0.69)  -13.0 (-17.4, -9.09) | -2.27 (-3.34, -1.28)  <0.01 (0.01, 0.01)  -13.1 (-17.5, -9.17) | -1.31 (-1.97, -0.75)  <0.01 (0.01, 0.01)  -12.3 (-16.4, -8.53) |
| Asthma  **Musculo-skeletal** | -781 (-1191, -420) | -635 (-965, -340) | -703 (-1069, -378) | -633 (-958, -343) | -615 (-945, -324) | -856 (-1329, -443) |
| Low back pain | -641 (-1272, 1.59) | -614 (-1216, -0.39) | -639 (-1248, -12.2) | -628 (-1210, -27.0) | -619 (-1258, 31.1) | -620 (-1295, 53.0) |
| Hip osteoarthritis  Knee osteoarthritis | 0.26 (0.26, 0.53  2.38 (1.85, 2.91) | 0.39 (0.39, 0.39)  1.95 (1.17, 2.35) | 0.36 (0.29, 0.43)  2.16 (1.65, 2.66) | 0.34 (0.28, 0.41)  1.93 (1.45, 2.34) | 0.28 (0.21, 0.36)  1.78 (1.35, 2.20) | 0.19 (0.09, 0.19)  0.94 (0.75, 1.22) |

Table S5: Change in incident cases of disease per 100,000 adult population (2018 to 2040), in specified local authorities as a result of the intervention, assuming 100% stringency
